# Genomic Epidemiology of Coxsackievirus A24 Variant During the 2024 Acute Hemorrhagic Conjunctivitis Outbreak in Coastal Kenya

**DOI:** 10.64898/2025.12.18.25342382

**Authors:** John Mwita Morobe, Edidah M. Ong’era, Arnold W. Lambisia, Samuel O. Odoyo, Martin Mutunga, Esther N. Katama, Robinson Cheruiyot, Joyce U. Nyiro, Everlyn Kamau, Charlotte J. Houldcroft, Matt Keeling, Katherine Gallagher, Edward C. Holmes, Charles N. Agoti

## Abstract

Several African countries experienced a surge in acute hemorrhagic conjunctivitis (AHC) cases in 2024. Investigations in Kenya, Mayotte and Tanzania identified coxsackievirus A24 variant (CV-A24v) as the causative agent. To date, limited genomic data exist to elucidate the sources, epidemiology, and evolution of CV-A24v in Africa. We generated 245 CV-A24v genomes from samples collected between January and September 2024 in coastal Kenya. Phylogenetic analysis showed that these viruses belonged to genotype IV, with two major clusters identified that differed by 52 nucleotides and five amino acids, with recombination detected in the 3D*^pol^* gene. The sequences clustered closely with contemporaneous Mayotte and Malawi sequences but were distinct from Asian sequences from 2023. Molecular clock dating revealed that the Kenyan sequences had a Most Recent Common Ancestor (MRCA) between June and October 2023. Our study provides the first detailed genomic analysis of CV-A24v from Africa to inform its future control strategies.

## Introduction

Coxsackievirus A24 variant (CV-A24v), a member of species *Enterovirus C*, genus *Enterovirus*, is a leading cause of acute hemorrhagic conjunctivitis (AHC)^1–4^, frequently occurring as large and explosive outbreaks in tropical and sub-tropical regions^5^. CV-A24v infection is characterized by rapid onset of ocular pain, sub-conjunctival hemorrhage, swollen eyelids, foreign body sensation, epiphora, eye discharge and photophobia^6–8^. Typically, the symptoms follow a short incubation period of 12-48 hours and persist for three-five days before resolving within 10-14 days^7,9^. CV-A24v infection is detectable in respiratory, saliva, stool and ocular samples^10–12^. Outbreaks drive substantial morbidity, productivity losses, and work/school absenteeism, increased visits to eye clinics, and often lead to unnecessary antibiotic use, contributing to antimicrobial resistance concerns^13–15^.

Since its discovery in 1970^16^, CV-A24v has been implicated in several outbreaks, especially in parts of Asia^3,4,17–19^, Africa^20–23^, and South America^24,25^, often affecting thousands of people within a short period of time^5^. CV-A24v outbreaks re-emerge at variable intervals^5^, typically 3-5 years apart^5^, with large-scale epidemics occurring approximately at 8-10 years intervals in tropical and subtropical countries^5^. How CV-A24v persists between outbreaks is not well understood. The re-emergence of CV-A24v in communities may be in part due to its antigenic evolution in the viral capsid (VP) proteins, as well as waning population immunity, both of which aid reinfections^24,26,27^. Neutralizing antibody immune responses have been found to decline significantly within one-year post-infection^28^.

Detailed molecular epidemiological investigations of CV-A24v in Africa remain scarce. This, in turn, has inhibited our understanding of its underlying evolutionary and transmission dynamics. For example, in Kenya, the characterisation of previous outbreaks has been limited to case studies or outbreak reports ^29^. In early 2024, an outbreak of AHC caused by CV-A24v was observed along the Kenyan coast^30,31^ coinciding with contemporaneous outbreaks from several other African countries including Tanzania, Rwanda, Burundi, Malawi, Uganda ^20,21,32^ Madagascar and Mayotte^22,23^. Coincidentally, CV-A24v outbreaks were reported across parts of Asia in 2023, including in Pakistan, China, India, Nepal, and Bhutan^1,3,17,18^. The geographic and temporal overlap or succession of these outbreaks suggests possible epidemiological linkage. However detailed CV-A24v.phylogenetic analyses to elucidate virus origins, genetic relationships, transmission pathways are lacking.

Here, we analyzed respiratory and stool samples collected through ongoing acute respiratory infection and acute diarrhea in Kilifi county, coastal Kenya, alongside ocular samples collected during the AHC outbreak in 2024 in Mombasa county^30,31^. A subset of positive samples underwent whole-genome sequencing(WGS) followed by combined phylogenetic and phylodynamic analysis with all publicly available CV-A24v genomes (n=159, as of 01 November 2025). This work builds on prior Kenyan^30,31^ and regional studies^20,23^, which largely focused descriptive epidemiology of previous CV-A24v detections. In doing so, we provide the first comprehensive genomic analysis of a CV-A24v outbreak in an African setting, detailing its genetic diversity, evolutionary patterns, and local transmission dynamics.

## Results

### CV-A24v detections in Kilifi

The distribution of samples collected, tested, CV-A24v positive, and sequenced across our Kilifi surveillance platforms is summarized in **Table 1**. The analysis identified the first CV-A24v positive sample from a ResViRe (<u>Res</u>piratory <u>Vi</u>rus <u>Re</u>infections) study participant on 23^rd^ January 2024. Thereafter, a rapid increase in CV-A24v detections was observed in the other surveillance platforms **(Figure 1).** Peak CV-A24v was observed in February 2024 for both ResViRe (7.1%), and the out-patient health facility (HF) (5.3%) studies, but in March 2024 in the paediatric in-patient (IP) pneumonia study:13.6% and diarrhoea study: 37.9%, **Figure 1).** Following this, the cases declined steadily decline until June 2024, with sporadic detections through to December 2024 **(Figure 1**).

**Figure 1.**
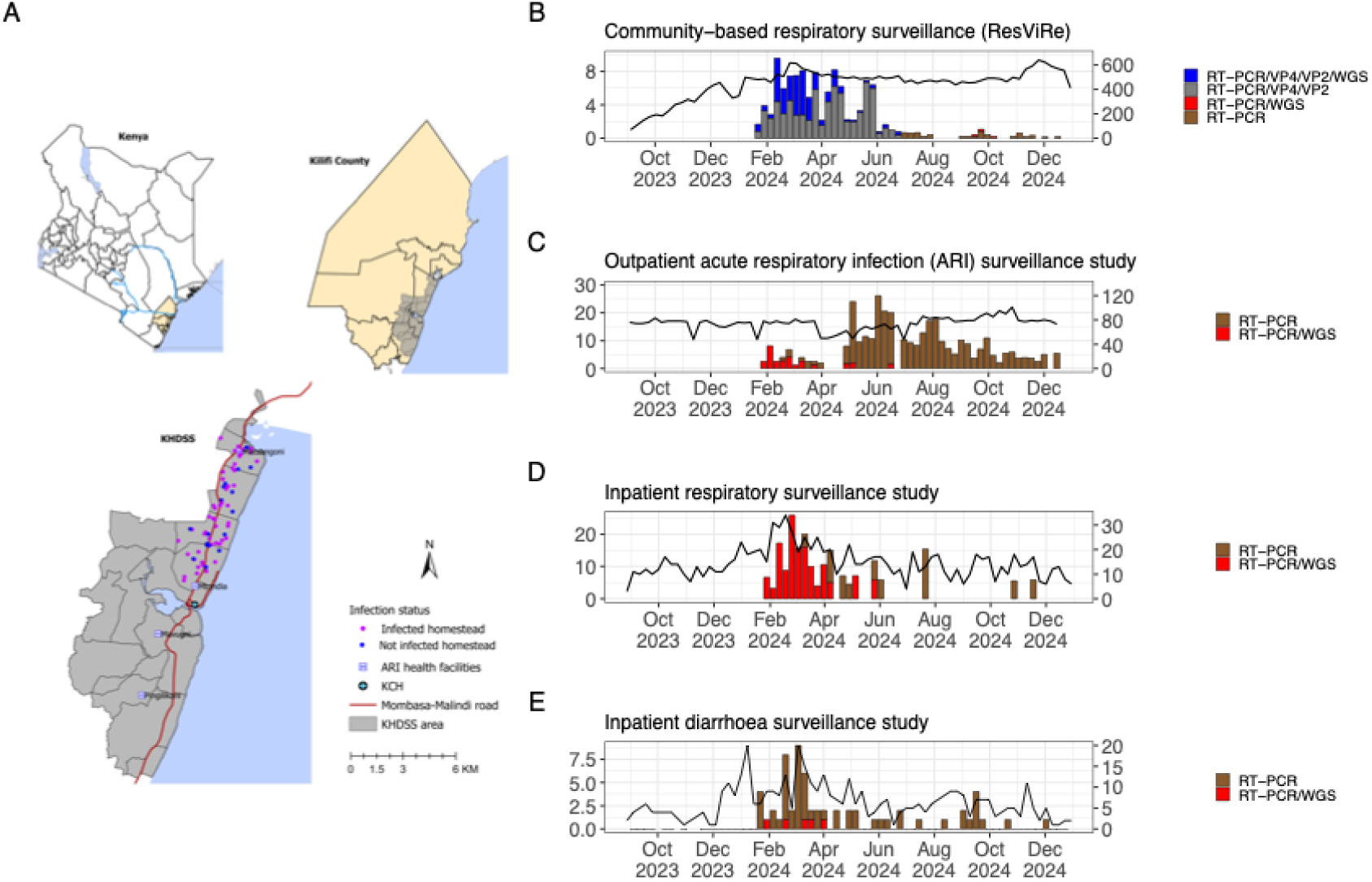
Study area and temporal distribution of CV-A24v positivity across the four surveillance platforms. (**A**) Map of Kenya, showing location of Kilifi county and KHDSS. The pink and blue dots show the distribution of the recruited homestead in the community surveillance arm. Location of the five health facilities for in-patient (KCH) and/or out-patient surveillance are indicated. (**B**) Temporal distribution of CV-A24v positives at the community-based respiratory surveillance. (**C**) Temporal distribution of CV-A24v positives at the in-patient pneumonia surveillance. (**D**) Temporal distribution of CV-A24v positives at the out-patient acute respiratory infection surveillance. (**E**) Temporal distribution of CV-A24v positives at the in-patient diarrheal etiology surveillance. The bar plot shows the percentage of samples positive for CV-A24v, stratified by the detection or confirmatory methods used, while the black line represents the total number of samples tested.

**Table 1.** Distribution of samples collected from AHC outbreak in Mombasa, and the various surveillance platforms in the KHDSS between August 2023 and December 2024, indicating the total number of samples collected, tested, CV-A24v positives, and successfully sequenced.

| Surveillance Platform | Period | Number of Samples | CV-A24v positive Samples RT-PCR | CV-A24v Positive Samples VP4/2 Sequencing | Samples Sequenced (WGS) |
| --- | --- | --- | --- | --- | --- |
| Community-based homestead surveillance study (ResViRe study) ‡ | 31-Aug-2023 – 31-Dec-2024 | 31,298 | 0 | 599 | 182 |
| Pediatric in-patient pneumonia surveillance | 01-Aug-2023 – 31-Dec-2024 | 1,088 | 48 | 0 | 33 |
| Health facility out-patient surveillance | 01-Jan-2024 – 31-May-2024 | 1,486 | 66 | 0 | 22 |
| In-patient diarrhea etiology surveillance | 01-Jan-2023 – 31-Dec 2024 | 391 | 64 | 0 | 5 |
| Mombasa AHC outbreak* | 15 February 2024 | 15 | - | 3 | 3 |
| **Total** | **-** | **34,301** | **178** | **599** | **245** |

‡In the ResViRe study, samples were collected from 713 individuals, of whom 218 (26.7%) tested positive for CV-A24v. Of these, 182 genome sequences were recovered, representing samples from 78 individuals.\*CV-A24v identified using metagenomic sequencing

CV-A24v-positive individuals were younger in the ResViRe study (median age 5 vs 19 years in negative individuals; p < 0.001), with over half (51%) of infections detected in 0–5 years (**supplementary Table 1**). Similarly, CV-A24v-positive participants were younger (median age 9 years vs 16 years; p = 0.004) in the out-patient surveillance, with children <5 years comprising 45% of positives compared to 29% of negatives. Detections in paediatric in-patient pneumonia and diarrheal admissions, were more frequently in infants (0–11 months). Significant differences were observed in positivity across administrative locations, with the Roka and Zowerani localities (p = 0.015) showing high positivity rates in the ResViRe study, and prevalence similarly varied by health facility (p = 0.042) in the HF study, being highest in Pingilikani and Mavueni. Positivity across by gender was comparable in all surveillance platforms (**supplementary Table 1)**.

### CV-A24v sequencing and genotyping

Whole-genome sequencing was attempted on 333 CV-A24v–positive samples collected between 23^rd^ January and 23^rd^ September 2024. Priority was given to samples with a qPCR cycle threshold (Ct) values <30.0 due to higher likelihood of sequencing success^33^ **(Table 1).** A total of 245 (74.0%) yielded genomes (coverage >98.0%). These ranged from 7,304 to 7,430 nucleotides (nt) in length and comprised a 5′ untranslated region (UTR) of ∼750 nt, a 3′ UTR of ∼69 nt, and a single open reading frame (ORF) of 6,645 nt.

Of the 245 genomes, 182 (74%) were generated from 78 individuals enrolled in the community-based household surveillance platform, of whom 43 contributed more than one sequence. For the remainder, 33 (13%) were from the in-patient pneumonia surveillance, 22 (9%) from the out-patient ARI surveillance, 5 (2%) from the diarrhoea aetiology surveillance and 3 (1%) from AHC outbreak in Mombasa county **(Table 1).** To date eight genotypes of CV-A24v have been identified from phylogenetic analyses to have circulated globally between 1970 to the present^34^. Both phylogenetic analysis and the Enterovirus Genotyping Tool (based on partial viral protein 1 (VP1) sequence)^35^ determined that all the 245 Kenyan sequences belonged to genotype IV.

### Global context of Kenya CV-A24v strains

Global phylogenetic analysis revealed viruses from the same continent and outbreak often formed monophyletic clusters **(Figure 2A and supplementary Figure 1).** The observed diversity within genotype IV warranted additional analyses to fully characterize the recent global CV-A24v circulation. On applying a clustering analysis to the VP1 coding region of genotype IV strains using VSEARCH v2.30.2^36^, sequences were dereplicated and clustered with the `--cluster_fast` algorithm at a 95% nucleotide identity threshold (corresponding to 0.05 divergence)^34^. This approach resolved genotype IV sequences into 22 distinct sub-groups (here in denoted as GIV-C1–C22) (**supplementary Figure 2b**). Notably, although all the 2024 Kenyan and other Eastern/Southern Africa sequences fell into the same sub-group, they separated into three clusters (herein designated as C1/24, C2/24, and C3/24) **(Figure 2 and supplementary Figure 1)**. The Kenyan strains from 2010 and 2024 AHC outbreaks fell into different subgroups. Global CV-A24v analysis showed that the 2024 Kenyan, Malawi, and Mayotte sequences belong to a different sub-group from the 2023 Asian strains associated with outbreaks in India, Nepal, Bhutan, China, and Pakistan. The 2024 African cluster, however, was more closely related to strains from the 2017–2018 AHC outbreaks in Uganda, French Guiana, Brazil, Cameroon, and Mexico **(supplementary Figure 1).**

**Figure 2.**
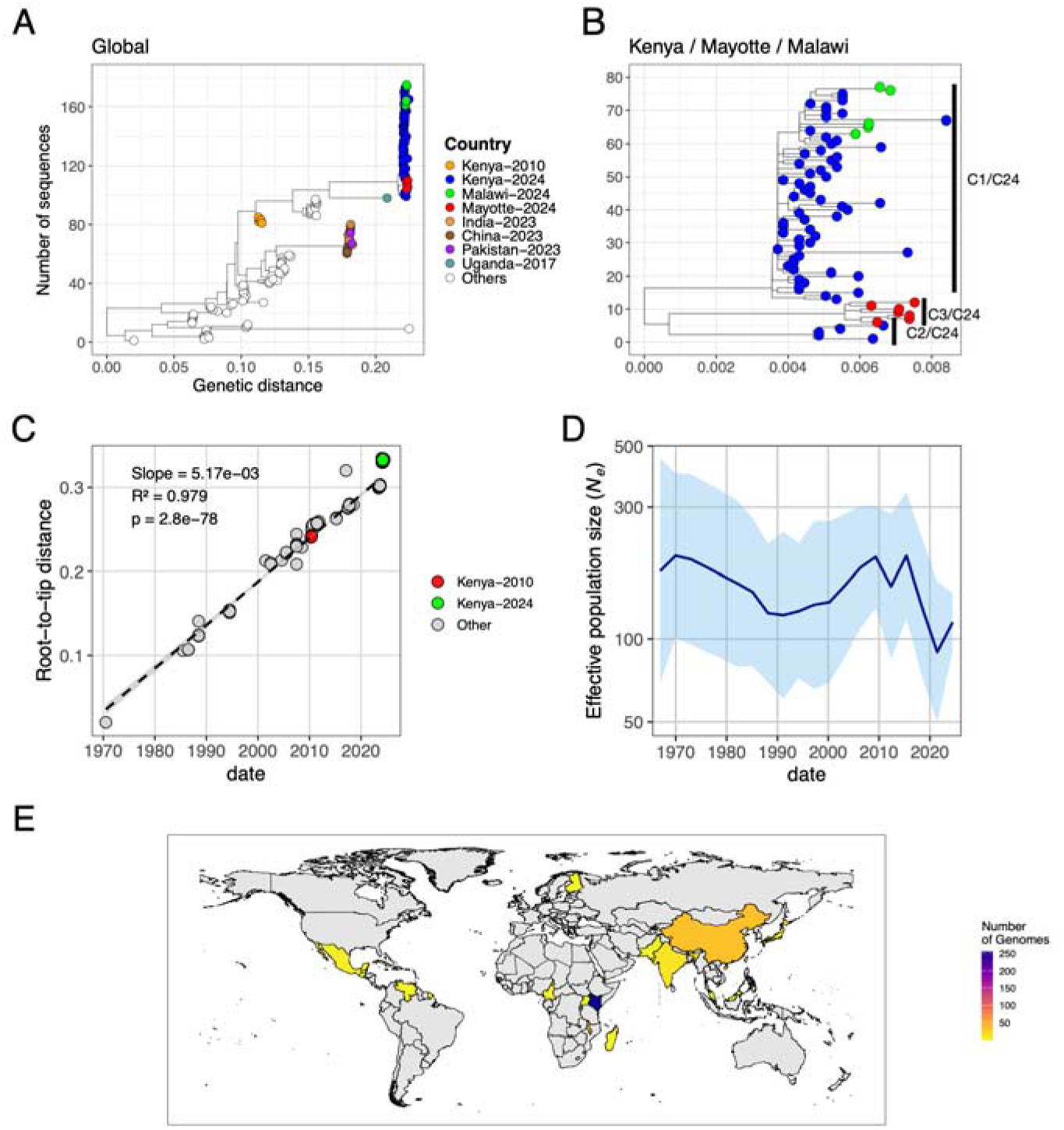
Genetic diversity, root-to-tip divergence, and demographic reconstruction of global CV-A24v strains sampled between 1970 and 2024 (**A**) ML phylogenetic tree of 194 whole genome sequences selected from the global (129) and Kenya (n=65) data sets. (**B**) ML phylogenetic tree of 80 sequences selected from Kenya, Mayotte and Malawi. Tip labels are colored by country of sampling, highlighting Kenya, Mayotte, and Malawi. (**C**) Root-to-tip regression showing accumulation of genetic divergence in CV-A24v over time. **(D)** ML skyline reconstruction of genetic diversity (presented as the effective population size, *Ne*) of CV-A24v over time, assuming 50 generations per year. The solid line represents the median estimate of *Ne*, while the shaded area shows the approximate 95% confidence bounds (±2 standard deviations of the log-likelihood). (**E**) World map showing the distribution of countries that have deposited CV-A24v WGS in GenBank as of 01 November 2025. Color intensity indicates the number of sequences contributed by each country.

### Timing of global CV-A24v MRCA and population size changes

The global CV-A24v data set showed a strong molecular clock signal from our root-to-tip regression analysis on the ML phylogeny (R^2^=0.975) **(Figure 2C).** The estimated mean evolutionary rate of the global sequences was 5.37 × 10^−3^ substitutions per site per year (95% HPD: 4.923 × 10^−3^ – 7.559 x 10^-3^ **(Figure 2C)**. Based on this rate, the tMRCA was estimated around December 1967 (95% HPD February 1961 – June 1970). Skyline coalescent model was used to infer the global demographic history of CV-A24v. This revealed marked variation in genetic diversity (reflected in effective population size (*Ne*) over time) that corresponded to outbreaks in different global regions: 1990–2000 in Asia, 2000–2010 in Asia and Africa, and 2015–2020 in South America, Africa and Asia **(Figure 2D)**. A sharp increase in *Ne* was observed from between 2022 and 2024, coinciding with the inclusion of sequences from recent outbreaks in Africa and Asia **(Figure 2D)**.

### Genetic diversity of Kenya and other Africa CV-A24v strains

Focusing on the local dynamics of the 2024 AHC outbreak, the C1/24 cluster was predominantly comprised of Kenya genomes with Malawi sequences interspersed within the cluster, the C2/24 cluster consisted exclusively of Kenya virus genomes, while the C3/24 cluster comprised exclusively sequences from the French Overseas Department of Mayotte **(Figure 3A).** The two identified clusters in Kenya co-circulated throughout the AHC outbreak (**Figure 3F)**. Within C1/24 and C2/24 multiple well-supported sub-clusters (bootstrap support >70) were observed, frequently including viruses from the same location or surveillance platform **(supplementary Figure 3**). The mean evolutionary rate of the 2024 Kenya genomes was estimated at 5.351 × 10⁻³ substitutions/site/year (95% HPD interval: 3.914 × 10⁻³ – 6.793 × 10⁻³) and the estimated tMRCA was around September 2023 (95% HPD interval : June 2023 – October 2023). Overall, the C1/24, C2/24 and C3/24 nucleotide sequence similarities ranged from 98.42% to 100% and amino acid similarities from 99.99% to 100%. Within clade C1/24, nucleotide similarities ranged from 99.10% to 100% and amino acid similarities from 99.6% to 100%, with equivalent values observed within the C2/24 and C3/24 clades. The pairwise single nucleotide polymorphism (SNP) differences among all sequences in the three clusters combined ranged from 0 to 101 SNPs (median: 76). Within cluster C1/24, SNP distances ranged from 0 to 52 (median: 17), in C2/24 they ranged from 8 to 33 (median: 12), whereas within cluster C3/24 they ranged from 8 to 25 (median: 14) **(Figure 3B-D).**

**Figure 3.**
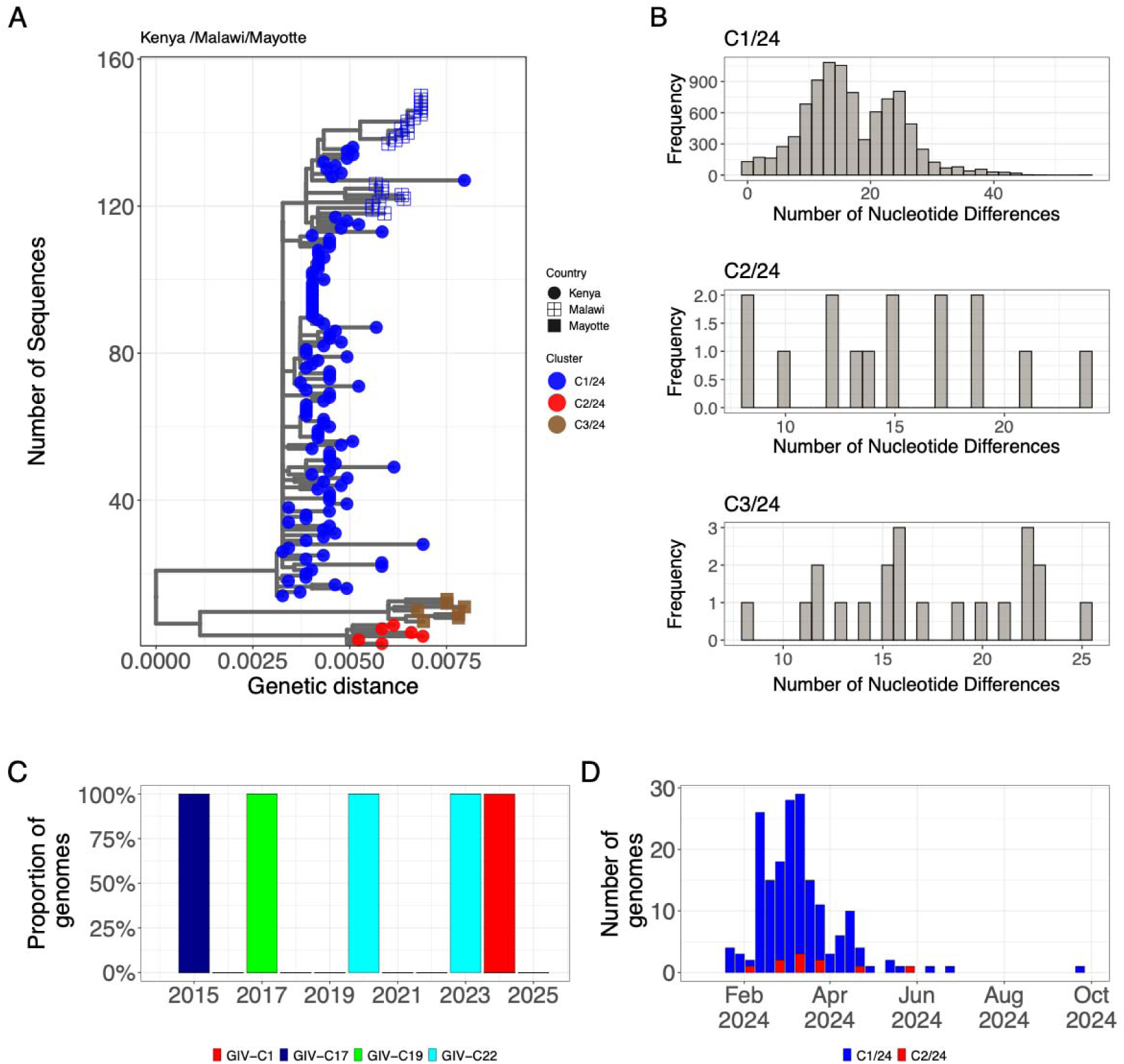
Genetic diversity of selected Kenya, Malawi and Mayotte CV-A24v sequences. (**A**) ML phylogenetic tree of the Kenya 2024 CV-A24v strains. Tips are colored according to major clade identified (C1/24 in blue, C2/24 in red and C2/24 in tan). Different shapes indicate country of sequence. (**B**) Pairwise SNP-distance distribution plots of C1/24, C2/24 and C3/24 sequences. (**C**) Temporal distribution of CV-A24v sub-groups detected between 2015–2024, classified using the established 0.05 VP1 pairwise divergence threshold for genotype IV (GIV) sub-group delineation. (**D**) Temporal distribution of the C1/24 and C2/24 clusters in coastal Kenya.

### Analysis of amino acid variation

The 2024 Kenya CV-A24v sequences were compared to the prototype strain EH24/70 (GenBank accession no. D90457), revealing amino acid substitution at 89–95 codon sites **(Figure 4A),** and 41–48 amino acid differences compared to the earliest genotype IV genomes. Most substitution occurred in VP1, 2C, and 3D, with each gene exhibiting substitutions at 15 codon sites **(Figure 4A).** The C1/24, C2/24 clusters were distinguished by five amino acid substitutions at codon sites 935 (I/T), 1371 (N/S), 1547 (H/Y), 2015(I/V) and 2213(L/S) **(Figure 4B).** Kenya 2024 CV-A24v sequences exhibited one amino-acid difference relative to Malawi strains and five when compared to Mayotte **(Figure 4B**), but diverged at 51 codon sites from 2023 Asia strains. Kenya 2024 CV-A24v sequences differed at 27 sites from Kenya 2010 sequences **(Supplementary Figure 4)**. Entropy analysis of the global CV-A24v dataset revealed substantial heterogeneity throughout the polyprotein. This pattern was mirrored in the Kenya-only dataset, where similarly elevated entropy values were observed along the genome **(Figure 4C and D).** Outside the protein coding region, the Kenyan strains exhibited a single-nucleotide insertion at position 102, a single-nucleotide deletion at position 585, and a four-nucleotide deletion spanning positions 117–120 within the 5′ untranslated region (UTR) relative to the prototype strain EH24/70.

**Figure 4.**
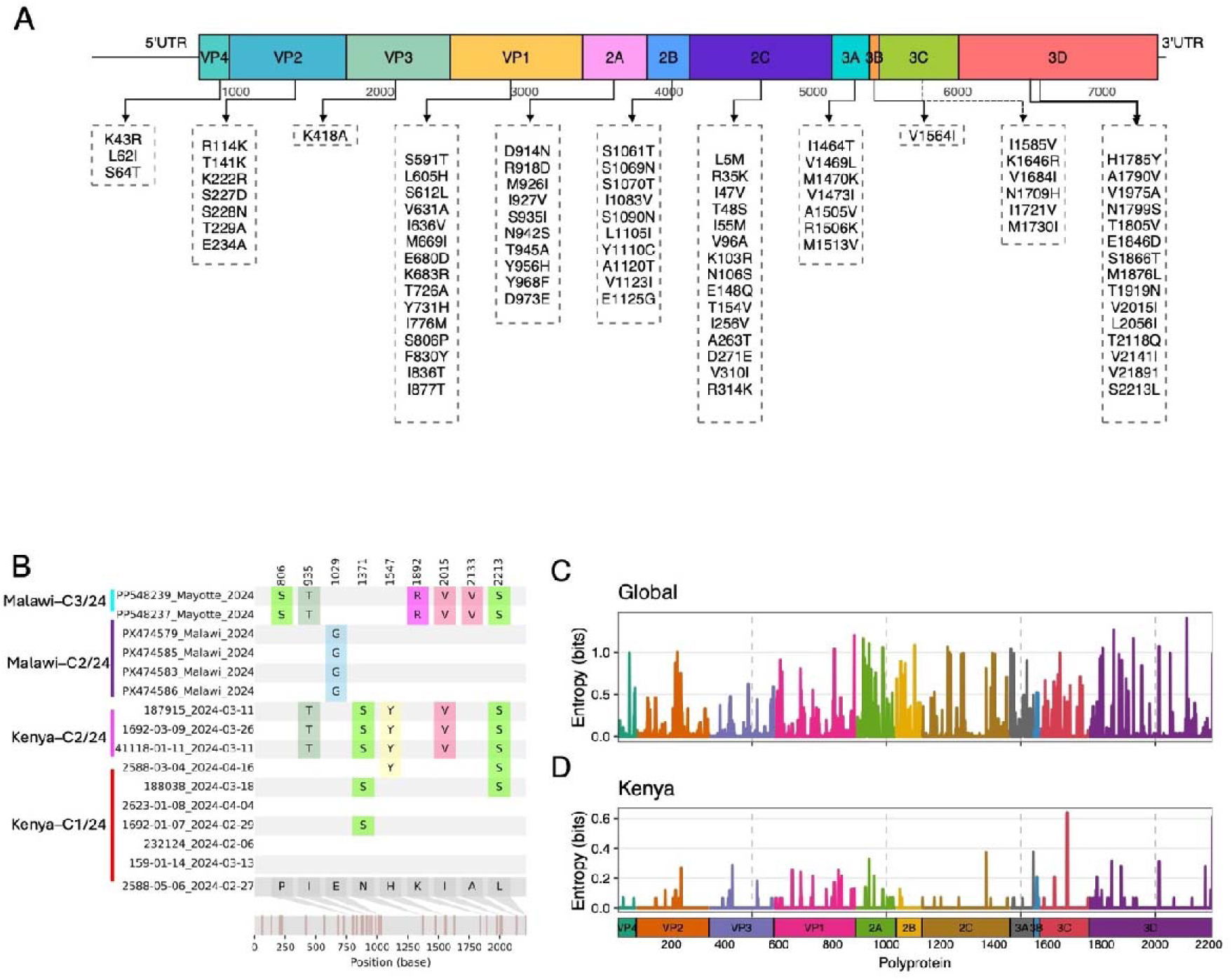
Amino acid variability across the CV-A24v genome. (**A**) Summary of amino acid changes across different genomic regions of the Kenya CV-A24v, comparing the prototype strain (D90457.1/Singapore/1970). (**B**) Amino acid differences between sequences from Kenya, Malawi, and Mayotte, with the Kenyan sequence acting as the reference. (**C**) Global sequences showing variable amino acid positions and their distribution across coding regions. **(D)** Kenyan sequences showing variable amino acid positions and their distribution across coding regions. Peaks represent regions with higher amino acid variability

Selection analyses using SLAC estimated a mean ratio of non-synonymous to synonymous substitutions (i.e., d_N_/d_S_) at 0.0706, indicative of strong purifying selection. Despite this, SLAC, FEL, and FUBAR collectively identified evidence of positive selection at 10 codon positions (**Appendix 1**). Under SLAC, five codons position – VP2 (234, 236), VP1 (683, 881), (2C) 1371 and (3C) 1601 showed evidence of increased fixation of non-synonymous mutations (d_N_/d_S_ >1). The FEL method identified 10 codon positions subject to positive selection: VP2 (234, 236), VP1 (683, 830, 871, 881), 2C (1371), 3A (1506), 3C (1601), 3D (2124). FUBAR also identified six codon positions under pervasive positive selection with posterior probabilities ≥ 0.9: codons VP1 (683, 830, 871, 881), 2C(1371), and 3C(1601) **(Table 2).**

**Table 2.** Codon sites inferred to be under positive selection in CV-A24v using SLAC, FEL, and FUBAR.

| Gene | Codon position | SLAC: dN–dS | SLAC p-value | FEL: $\alpha$ (dS) | FEL: $\beta$ (dN) | FEL p-value | FUBAR: $\alpha$ | FUBAR: $\beta$ | FUBAR posterior ($\beta > \alpha$) |
| --- | --- | --- | --- | --- | --- | --- | --- | --- | --- |
| VP2 | 234 | 3.955 | 0.043 | 0 | 1.434 | 0.0164 | – | – | – |
| VP2 | 236 | 3 | 0.088 | 0 | 0.996 | 0.0396 | – | – | – |
| VP1 | 683 | 6.046 | 0.041 | 0.418 | 2.166 | 0.0598 | 0.994 | 6.782 | 0.948 |
| VP1 | 830 | – | – | 0.642 | 2.562 | 0.0537 | 1.008 | 6.801 | 0.949 |
| VP1 | 871 | – | – | 0.321 | 1.645 | 0.0735 | 0.843 | 5.278 | 0.933 |
| VP1 | 881 | 5.794 | 0.055 | 0.311 | 2.887 | 0.0063 | 0.677 | 8.286 | 0.998 |
| 2C | 1371 | 4.342 | 0.036 | 0 | 1.895 | 0.0029 | 0.441 | 6.448 | 0.995 |
| 3A | 1506 | – | – | 0 | 1.07 | 0.0453 | – | – | – |
| 3C | 1601 | 5.218 | 0.089 | 0.395 | 2.29 | 0.0461 | 0.828 | 6.422 | 0.959 |
| 3D<sup>pol</sup> | 2124 | – | – | 0 | 0.809 | 0.0742 | – | – | – |

Codons were considered to be under strong positive selection if supported by at least two independent methods, defined as SLAC $p < 0.05$ , FEL $p < 0.05$ , or FUBAR posterior probability $\geq 0.9$ for $\beta > \alpha$ .

### Recombination of CV-A24v with other enteroviruses

Similarity analysis of the CV-A24v strains that circulated in East/Southern Africa in 2024 and globally using SimPlot++^37^ suggested that the 2024 Kenya/Mayotte/Malawi viruses may have a recombinant history**(Figure 5A).** The breakpoint for the putative recombination event was identified to be located within the 3D*^pol^* genome region **(Figure 5).** To identify potential parental/minor strains, a BLAST analysis of the 3D*^pol^* genome region was conducted using the GenBank database as of 01 November 2025. The 3D*^pol^* genome region sequences from the 2024 Kenya/Mayotte/Malawi strains showed between 70 and 88% similarity when compared to other enteroviruses (EVs) sequences assigned to the *coxsackiepol* species (i.e. poliovirus type 1, 2 and 3). Recombination analysis using multiple programs within the Recombination Detection Program (RDP v5.74) confirmed recombination within the 3D*^pol^* region of sequences from Kenya, Malawi and Mayotte, and the 2017 Uganda strain. The event involved CV-A24v as the major parental sequence and Poliovirus type 1 as the minor parent (**Figure 5B**). Phylogenetic trees were constructed for sequences on either side of the breakpoint alongside poliovirus types 1–3, revealed discordant clustering topologies **(Figure 5C and D**). Across coding regions, CV-A24v sequences clustered together and remained distinct from polioviruses, indicating largely consistent topologies and limited intra-species recombination. An exception was the 3D*^pol^* region, where 2024 Kenya/Mayotte/Malawi (2024) and Uganda (2017) sequences clustered closer to polioviruses, suggesting a recombination event **(Supplementary Figure 5).**

**Figure 5.**
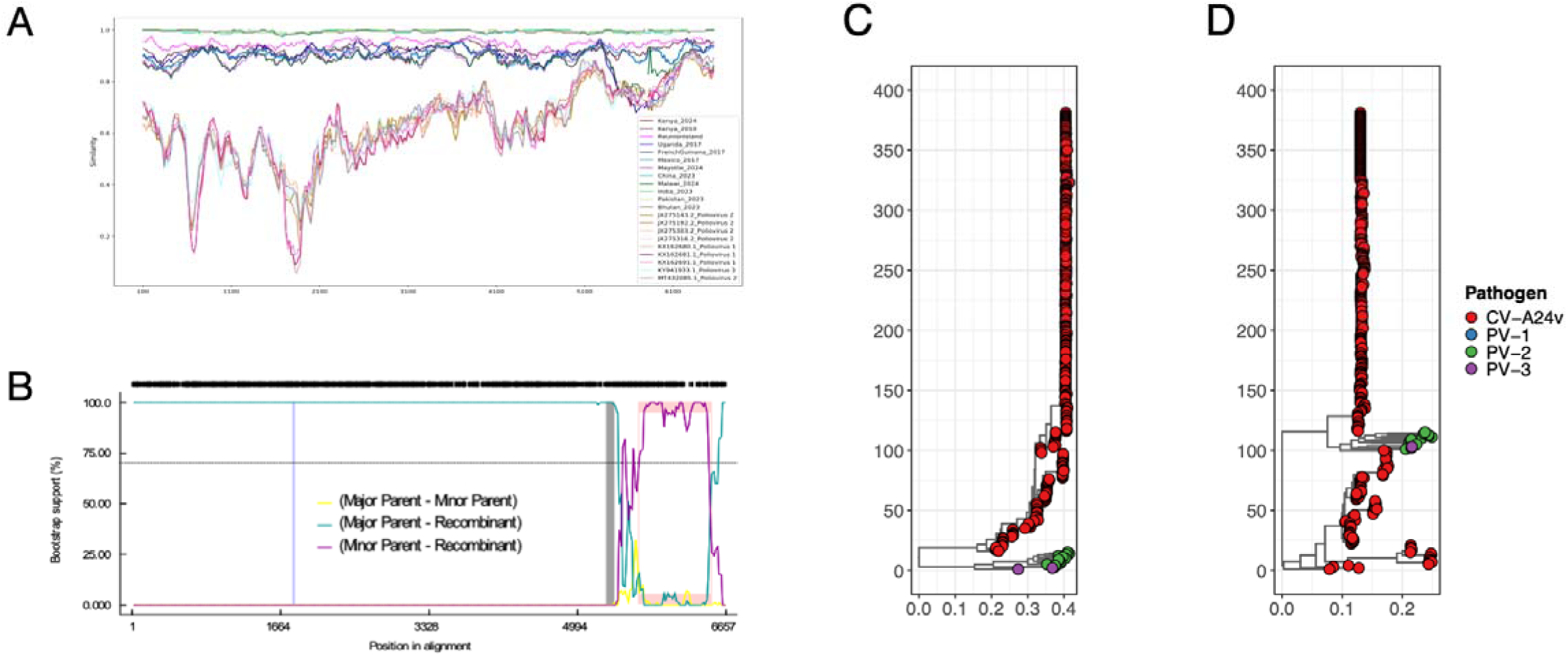
Nucleotide similarity between CV-A24v and other species C enteroviruses. **(A)** Nucleotide similarity plots between Kenyan CV-A24v genomes and other selected sequences from enterovirus C sequences obtained from GenBank as of 15th August 2025. **(B)** RDP5 BOOTSCAN analysis of the selected enterovirus C and CV-A24v strains. The analysis was based on a 200-nucleotide sliding window with a 20-nucleotide step size. **(C)** ML tree for CV-A24v sequences spanning nucleotide positions 1–5253, and **(D)** ML tree for sequences spanning positions 5253–7400.

## Discussion

We leveraged samples collected from ongoing respiratory/diarrheal disease surveillance within the Kilifi Health and Demographic Surveillance (KHDSS)^38^ area, Kilifi county, and during the AHC outbreak response activity in Mombasa county^30,31^, to produce the largest set ever of CV-A24v genomes from a single site in Africa and globally to date. Prior to this study, only 10 CV-A24v genomes from Kenya existed on GenBank. Our new data provide valuable insights into CV-A24v genomic epidemiology at the local scale, evolutionary dynamics, and regional epidemic inter-connectedness, especially the 2024 outbreak in East Africa.

In this study, CV-A24v cases were first detected on the 23^rd^ of January 2024 through community-based household surveillance, after which a rapid surge was observed in all surveillance platforms. The highest positive sample prevalence was observed in the in-patients diarrheal and respiratory platforms, which predominantly sample severely ill children aged <5 years. Notably, the community homestead-based household surveillance and the outpatient health facility surveillance that recruited patients across all age groups also observed a bias of infections towards the younger age groups. Together, this suggests that CV-A24v infections and disease burden is most pronounced in childhood, a pattern consistent with the epidemiological profile of many enterovirus infections^39,40^ and previous reports of CV-A24v -associated AHC outbreaks^19,41^.

The Kenya 2024 CV-A24v sequences fell within genotype IV, with two major clusters (C1/24 and C2/24) identified. Similar patterns have been seen in previous AHC outbreaks, where divergent clusters within genotype IV co-circulated during a single outbreak^34,42^, suggesting sustained community transmission and *in situ* diversification driven by both multiple introductions and local spread^10,24,43^. We observed close phylogenetic relationship between the 2024 Kenyan sequences and strains from Mayotte and Malawi strains, and earlier Uganda strains suggesting regional persistence of the virus for many years^20,23,30^. At the global scale, the 2023–2024 AHC outbreaks appear to be distinct by continent, with different genotype IV clades predominating different geographic regions. The viral clade associated with the 2023 Asia AHC outbreaks (Bhutan, Pakistan, India, China and Nepal)^3,17,18,43^ was distinct from that associated with the Kenya, Malawi, and the Indian Ocean islands (and Mayotte) outbreaks^20,21,23,30,32^.

Despite this genetic divergence at the global scale, both lineages drove large-scale, near-synchronous outbreaks. For this reason, viral evolution alone is unlikely to explain the parallel resurgence. Previous immunologic studies have demonstrated that immunity to CV-A24v declines rapidly after infection (approximately less than 1 year)^28^, leading to replenishment of new susceptibles in addition to births. Seasonal and environmental factors may also influence emergence of AHC outbreaks^44–46^. For instance, AHC incidence in China showed a strong correlation with warm and humid months^46^, implying that weather conditions enhance viral stability or transmission via fomites and ocular secretions. Similarly, the Kenyan outbreak occurred during warm, humid months, and a period marked by unusually strong winds. Hence, it is likely that multiple factors converge to spark a new CV-A24v epidemic: antigenic novelty, waning of previous immunity and environmental conditions that support virus particle survival and dispersal ^23,26^.

The viruses sampled in Kenya, Mayotte and Malawi in 2024 were highly similar and closely related to a virus from Uganda (2017). The Ugandan 2017 sequence had the recombination signal that was also present in the 2024 Kenya, Malawi and Mayotte sequences. This supports the notion of a recent shared ancestry of CV-A24v strains observed in East/Southern Africa. It is difficult to discern whether these viruses were cryptically transmitting in the region and hence undetected^23,31^ or were reseeded from unknown source. The nucleotide similarity of the viruses sampled within East Africa regions and the MRCA suggest that the CV-A24v strains may have been circulated for a few months first detection during the 2024 AHC outbreaks. The continued sporadic detection of positive cases in coastal Kenya after the peak of the outbreak support the hypothesis that CV-A24v may continue to circulate during inter-epidemic times albeit at low levels, remaining un-detected.

Currently, CV-A24v is classified into eight genotypes (G-I–VIII), although all recent studies reported detection of only genotype G-IV. Within genotype IV, we observed substantial genetic divergence, highlighting the need to update the current classification scheme to track emerging genotypes. We suggest a systematic re-evaluation of the current CV-A24v genotyping scheme to reflect its recent genomic epidemiology, with additional new genotypes/ sub-genotypes/lineage assigned. Furthermore, the detection of a recombination event among Kenya, Mayotte, Malawi and Uganda strains highlights the genomic plasticity of CV-A24v and its potential role in shaping viral evolution.

The observed accumulation of amino acid mutations relative to the prototype strain reflects ongoing evolution of CV-A24v since its discovery in 1970^24,47^. The CV-A24v sequences have shown an accumulation of changes throughout its genome rather than confined to a specific region^47^. As expected, CV-A24v evolution is dominated by strong purifying selection, with only a small number of codon position showing signature of adaptive change. Indeed, different approaches (FEL, SLAC, FUBAR) consistently detected a limited number of codon positions under positive selection, particularly codon sites in VP1 (683, 830, 871, 881), 2C (1371), and 3C (1601), underscoring their potential functional relevance and the localized adaptive pressure in otherwise highly conserved genomic regions. These positively selected sites merit further structural or phenotypic investigation.

Skyline coalescent analysis revealed marked temporal fluctuations in *Ne* over time. An increase in *Ne* was observed in the early 1970s, corresponding to the emergence and global spread of CV-A24v during the first recorded outbreaks of AHC in Singapore, China and Taiwan^8,16^. Subsequent peaks in *Ne* corresponded to documented epidemic waves in the mid-1980s (China and Thailand), early 2000s (Korea India and Philippines), late 2000s (China and Cambodia), and mid-2010s (mainly in Southeast Asia and South America. Each of these expansion phases was followed by sharp declines in *Ne*, consistent with inter-epidemic periods characterized by reduced transmission and limited genomic sampling. The most recent rise in *Ne* during 2023–2024 coincided with widespread CV-A24v activity across Eastern/Southern Africa, the Indian Ocean islands, and South-East Asia. Such phylodynamic patterns are consistent with CV-A24v’s epidemiological signature of short, explosive outbreaks interspersed with prolonged periods of non-detection.^23,26^

This study has limitations. First, almost all participants in the study were recruited with a goal of identifying viral acute respiratory infection or diarrheal illness disease agents but not AHC. For instance, symptoms consistent with AHC were not systematically documented and thus clinical epidemiology of identified strains cannot be properly investigated here. Consequently, the accurate CV-A24v epidemiological patterns of this infection at the population level might require future focused investigation of this virus/disease. Second, there was limited availability of representative CV-A24v genomic sequences from outbreaks in neighboring countries and across the region. This paucity of regional genomic data reduces the power to fully resolve transmission routes, infer viral introductions, or contextualize local viral diversity within broader Eastern/Southern African or global circulation patterns. Third, the CV-A24v RT-PCR primers used in this study can cross-react with other enteroviruses, E.g Coxsackievirus A13. As a result, some RT-PCR positives may not represent CV-A24v infections, highlighting the need for confirmatory partial or WGS to accurately identify the pathogen.

In conclusion, following our sequencing of a large set of CV-A24v sequences from coastal Kenya, we demonstrate that the 2024 AHC epidemic in Kenya was driven by two distinct CV-A24v clusters within genotype G-IV – C1/24 and C2/24 – that have experienced sporadic adaptive evolution. These lineages, that likely diverged around mid-2023, clustered closely with contemporaneous CV-A24v sequences sampled in East/Southern Africa and Indian Ocean Islands. However, these lineages were also genetically distinct from other G-IV clade viruses sampled in parts of Asia in 2023, demonstrating the value of genomic data in understanding epidemic interconnectedness.

## Methods

### Study design and population

The samples analysed in this study were collected during multiple studies in coastal Kenya of varied designs conducted by the Kenya Medical Research Institute–Wellcome Trust Research Programme. These included (i) disease outbreak response activities^30^, and (ii) participants enrolled in ongoing respiratory/diarrhoea disease surveillance with the KHDSS^38^ **(Figure 1A).** The details of the studies are provided below:

a. A community-based household surveillance study named ResViRe (Respiratory Virus Reinfections) study. This was initiated in August 2023 and was ongoing throughout 2024 across five administrative locations: Tezo, Roka, Zowerani, Matsangoni, and Ngerenya. Here, recruited household participants were visited weekly at home and their respiratory disease status, travel history recorded and nasopharyngeal and oropharyngeal (NP/OP) swabs taken regardless of symptom status. Whenever one or more household member(s) were found to be positive for severe acute respiratory syndrome coronavirus 2, respiratory syncytial virus or influenza A/B, sample collection frequency for the household was increased to twice a week until all members test negative. By 31st December 2024, 67 homesteads (713 participants) had been enrolled and sampled at least once.
b. Paediatric pneumonia in-patient (IP) surveillance study at Kilifi County Hospital (KCH). This has been ongoing since 2002^48^. Respiratory samples are collected from children aged <5-year-old admitted to KCH with clinical symptoms of severe or very severe pneumonia as per WHO guidelines^49^.
c. Health facility out-patient (OP) acute respiratory infection (ARI) surveillance study. This began in December 2020 and was ongoing throughout 2024^50^. The study was undertaken at five selected health facilities with the KHDSS: Matsangoni, Mavueni, Pingilikani, Mtondia and the KCH out-patient department. At each health facility up to 15 NP/OP swabs are collected each week from persons of all ages presenting with ARI.
d. The IP childhood diarrhoea aetiology (enteric pathogens) surveillance study. This began at KCH in September 2009 and was ongoing throughout 2024. The study was established to track rotavirus infections/disease burden in the KHDSS population to provide impact data following introduction of rotavirus vaccination programme. Stool samples are collected from all eligible children <13 years of age admitted to the paediatric ward with diarrhoea as one of their presenting symptoms.

### CV-A24v detection and partial sequencing

CV-A24v was identified in clinical samples using a quantitative reverse transcription polymerase chain reaction (qRT-PCR) and/or sequencing of the VP4/VP2 genomic region of the enterovirus genome. Samples collected through ResViRe study between August 2023 and June 2024 were initially screened for rhinoviruses using qRT-PCR assay but this assay happens to capable of detecting a range of enteroviruses including CV-A24v. The assay targets the 5′ untranslated region (5′UTR) using the following primers and probes; forward primer (TGGACAGGGTGTGAAGAGC, reverse primer (CAAAGTAGTCGGTCCCATCC) and probe (VIC-TCCTCCGGCCCCTGAATG-TAMRA). A negative control (NC) (nuclease-free water) and positive control (of known RNA copies of human rhinovirus) were included in the assay. Samples were considered positive if Ct value <= 35.0. VP4/VP2 genomic region in the positive samples were subsequently amplified using forward primer (CCGGCCCCTGAATGYGGCTAA) and reverse primer (TCWGGHARYTTCCAMCACCANCC) and sequenced on the Illumina MiSeq platform. Raw Illumina reads were quality-filtered (Phred score < 20) and adapter-trimmed using fastp, followed by *de novo* assembly with SPAdes. The contigs were mapped to a reference HRV genome before generation of consensus sequences.

Resultant VP4/VP2 sequences were then analyzed using the Enterovirus Genotyping Tool^35^ to assess the presence of CV-A24v in the samples. Samples collected after June 2024, as well as those from other studies, were screened using a CV-A24v specific qRT-PCR assay adapted from Lévêque et al^51^. The assay targets the 5′UTR and uses the following primers: forward primer 5′-CCAACCACGGAGCAGGTGA-3′, reverse primer 5′-GAAACACGGACACCCAAAGTAGT-3′, and probe 5′-CAACCCAGCAACTAGCCTGTCGTAACGC-3′.

### Whole genome sequencing of CV-A24v

Whole genome recovery from CV-A24v-positive samples was performed using a three amplicon-targeted sequencing protocol established in our laboratory^31^. Briefly, viral RNA was reverse transcribed using the LunaScript® RT SuperMix Kit (New England Biolabs). A negative control (NC) (nuclease-free water) was included during both the extraction and reverse transcription steps. The cDNA was then amplified in three reaction tubes with the Q5® Hot Start High-Fidelity 2Master Mix (NEB) with optimised CV-A24v primers and thermocycling conditions as previously described^31^. The PCR products were loaded on a 1.5% agarose gel to confirm amplification before purification using Agencourt AMPure XP beads. Library preparation was performed using the Ligation Sequencing Kit (SQK-LSK114) and Native Barcoding Kit (NBD96), and sequencing performed on the Oxford Nanopore Technologies (ONT) GridION platform.

### Genome assembly

Genome assembly was performed using a sub-workflow of an in-house bioinformatics pipeline named *ViralPhyl*v0.11.2 and available on GitHub (https://github.com/kwtrp-peo/viralphyl). Base-called reads were demultiplexed using the ARTIC Guppyplex tool with default parameters, applying a minimum Q score of 9. Reads shorter than 400 nt were filtered out using the toullingQC module. Consensus sequences were generated by aligning the reads to a reference sequence (in this case CVA24_2400060741_FRA24, GenBank accession PP548240). The reads were aligned using MiniMap2^52^. Positions with genome coverage below 20 reads were masked with ’N’. The resulting consensus sequences were further refined using Clair v 2.1.1^53^.

### Global comparison data set

All CV-A24v sequence available in GenBank as of 01 November 2025 and with >90 % coverage were retrieved from GenBank (https://www.ncbi.nlm.nih.gov/genbank/). This dataset was used to assess the genetic relatedness of Kenyan CV-A24v strains to global strains, identify potential geographic origins, and infer patterns of lineage emergence, persistence, or reintroduction in East Africa. In total,159 genomes sampled between 1970 and 2024 from 21 countries across Africa, Asia, the Americas, and island nations in the Pacific Oceans were included in the analysis (**Figure 2E)**.

### Sequence Analysis

All the retrieved global sequences were collated and aligned using MAFFT v7.520117^54^. Nucleotide (nt) and amino acid (aa) variations were subsequently calculated using MEGA v11.0^55^ and the `dist.dna()` function in the `ape`5.0^56^ in R. To identify amino acid mutations, the Kenyan sequences were compared against the prototype strain EH24/70 (GenBank accession no. D90457). The nt and aa variations were analysed and visualised in Snipit v1.618^57^.

### Phylogenetic Analysis

To investigate the global phylogenetic structure and evolutionary dynamics of CV-A24v, the Kenyan CV-A24v sequences were analyzed together with the 159 public genomes (>90% coverage) retrieved from GenBank (as of 01 November 2025) **(Appendix 2)**. To avoid over-representation of closely related sequences and accurately capture viral diversity, the Kenyan genomes (n=245) were sub-sampled to account for repeated sampling from the same individual, household, or homestead, where samples were likely to be epidemiologically linked. In such cases, a single representative genome typically the earliest sampled or highest-quality sequence was retained, resulting in 65 sequences for downstream phylogenetic and phylodynamic analyses.Sequence alignment was performed using MAFFT v7.520^54^ with the --auto strategy to optimize alignment parameters for nucleotide data. The aligned data set was manually inspected and trimmed in AliView v1.30^58^. A maximum likelihood (ML) phylogenetic tree of this alignment was then inferred using IQ-TREE v2.1.3 (http://www.iqtree.org/) under the General Time Reversible (GTR+Γ) nucleotide substitution model, as determined by ModelFinder^59^. Node support was evaluated using 1,000 ultrafast bootstrap replicates, and the resulting tree was visualised and annotated using FigTree v1.4.4^60^ and ggtree v3.16.3^61^ in R^62^.

### Phylodynamic Analysis

The temporal signal of the sequence dataset was first assessed using TempEst v1.5.3^63^ by performing a regression of root-to-tip genetic distances against sampling dates. Estimation of the evolutionary rate and Time to the Most Recent Common Ancestor (tMRCA) and nucleotide substitution rate was performed using the Bayesian Markov Chain Monte Carlo (MCMC) approach available in the Bayesian Evolutionary Analysis Sampling Tree (BEAST) software package (v1.10.4 31)^64^ as well as in TreeTime^65^. Bayesian MCMC analyses were run for 500 million generations, with sampling performed every 1000 generations. Convergence of parameters were assessed in Tracer v1.7.1 (http://tree.bio.ed.ac.uk/software/tracer/. Convergence was considered to have been achieved when the effective sample size (ESS) of key parameters. In all cases, the initial 10% of the run was used as “burn-in”. A Maximum Clade Credibility (MCC) tree was generated using TreeAnnotator v1.8.4, and the resulting phylogeny visualized in FigTree v1.4.4^60^ or ggtree v3.16.3^61^ in R^62^. To infer temporal changes in viral population size through time values of the effective population size (*Ne*) were estimated using a skyline coalescent model^65^.

### Natural selection analysis

Selection pressures across the coding region of all CV-A24v sequences, including Kenyan strains, were assessed using HyPhy 2.5^66^, applying multiple detection methods: Fixed Effects Likelihood (FEL), Single Likelihood Ancestor Counting (SLAC), and Fast, Unconstrained Bayesian Approximation (FUBAR). FEL was used to identify codons under pervasive positive (d_N_ > d_S_) and negative selection (d_N_ < d_S_). SLAC provided estimates of d_S_ and d_N_ across the alignment based on maximum likelihood ancestral reconstruction, while FUBAR estimated codon-specific differences between nonsynonymous and synonymous rates under a Bayesian framework. For SLAC and FEL, significance was defined as P < 0.1, whereas for FUBAR, a posterior probability ≥ 0.9 indicated evidence of selection.

### Recombination Analysis

Potential recombination events were investigated using a combination of the (Recombination Detection Program) RDP5^67^ and SimPlot++^37^. Aligned genomes were screened with multiple RDP5 algorithms, including RDP, GENECONV, BootScan, MaxChi, Chimaera, SiScan, and 3Seq, applying Bonferroni-corrected p-values < 0.05 to confirm recombination signals detected by at least three independent methods. Breakpoint were mapped relative to the CV-A24v reference genome, and parental lineages were inferred where possible. SimPlot analyses used a 200 bp sliding window and 20 bp step size to visualise breakpoints. To further assess the recombination with other *Enteroviruses C* strains, phylogenetic trees were constructed for regions on either side of identified breakpoints and topologies compared to assess consistency and detect potential recombination events.

## Acknowledgements

We thank study participants, parents and guardians who consented participation in the various studies, members of the Pathogen Epidemiology and Omics (PEO) Group at the Kenya Medical Research Institute (KEMRI)-Wellcome Trust Research Programme particularly the field team who collected the clinical samples and metadata, and laboratory diagnostics team who processed the samples analysed and presented in this report. We thank Mr. Christopher Nyundo for his assistance in generating the study area maps.

## Authors’ contributions

JM, EM, AWL, RC, and MM contributed to the investigation. SO developed the bioinformatics tool used to assemble CV-A24v genomes. CNA secured funding and conceptualized the study. JM and ENK contributed to data curation and visualization. CNA, ECH, KG, and JN provided supervision. JM and CNA drafted the original manuscript. EK, CJH, MK, KG, ECH, and CNA contributed to reviewing and editing the manuscript. All authors read and approved the final manuscript.

## Data availability

Two hundred and forty-five WGS of CVA24v generated in this study have been deposited in the GenBank under the accession numbers PX241976-PX242099 and PX659063-PX659177. Epidemiological data, code and scripts for data analysis are available on the Harvard Dataverse: DOI: https://doi.org/10.7910/DVN/K3MIQV

## Ethics approval

Each surveillance platform (community, outpatient, and in-patient) that pro-vided samples analyzed here had a dedicated research protocol. The protocols’ consenting and sample collection process were reviewed and approved by KEMRI Scientific Ethics and Research Unit (SERU), Nairobi Kenya (protocol numbers #3178, #3103 and 4724). Samples were collected following consent from a parent or guardian for participants aged18 years. In addition, three ocular samples analysed in this study were collected during the Ministry of Health’s AHC outbreak response, where written informed consent is not required in public health emergency settings; these were fully anonymized and sample processing approved for use by SERU under Protocol #KEMRI/SERU/CGMR-C/304/4894 (approved May 19th, 2024).

