## Supplementary Material for "Genomic Epidemiology of Coxsackievirus A24 Variant During the 2024 Acute Hemorrhagic Conjunctivitis Outbreak in Coastal Kenya"

**Supplementary Materials**

**Supplementary Table**

**Supplementary Table 1**. Demographic characteristics of participants across the different surveillance platforms.

|  |  | **ResViRe** | |  |  |
| --- | --- | --- | --- | --- | --- |
| **Characteristic** | **Overall N = 714** | **CV-A24v negative N = 496** | | **CV-A24v positive N = 218** | **p-value** |
| Sex |  |  | |  | 0.3 |
| Female | 408 (57%) | 290 (58%) | | 118 (54%) |  |
| Male | 306 (43%) | 206 (42%) | | 100 (46%) |  |
| Median Age (years) | 14 (4, 29) | 19 (8, 35) | | 5 (1, 12) | <0.001 |
| Age Category (Years) |  |  | |  | <0.001 |
| 0-5 | 207 (29%) | 95 (19%) | | 112 (51%) |  |
| 6-10 | 97 (14%) | 55 (11%) | | 42 (19%) |  |
| 11-20 | 143 (20%) | 115 (23%) | | 28 (13%) |  |
| 21-30 | 94 (13%) | 78 (16%) | | 16 (7.3%) |  |
| 31-40 | 77 (11%) | 68 (14%) | | 9 (4.1%) |  |
| 41-50 | 34 (4.8%) | 32 (6.5%) | | 2 (0.9%) |  |
| >50 | 62 (8.7%) | 53 (11%) | | 9 (4.1%) |  |
| Location |  |  | |  | 0.015 |
| Matsangoni | 132 (18%) | 93 (19%) | | 39 (18%) |  |
| Ngerenya | 144 (20%) | 103 (21%) | | 41 (19%) |  |
| Roka | 141 (20%) | 87 (18%) | | 54 (25%) |  |
| Tezo | 151 (21%) | 119 (24%) | | 32 (15%) |  |
| Zowerani | 146 (20%) | 94 (19%) | | 52 (24%) |  |
| **Health facility out-patient surveillance** | | | | | |
| **Characteristic** | **Overall N = 1,486** | **negative N = 1,420** | | **positive  N = 66** | **p-value** |
| Sex |  |  | |  | 0.087 |
| Female | 871 (59%) | 839 (59%) | | 32 (48%) |  |
| Male | 615 (41%) | 581 (41%) | | 34 (52%) |  |
| Median Age (years) | 16 (4, 30) | 16 (4, 31) | | 9 (2, 18) | 0.004 |
| Age Category (Years) |  |  | |  |  |
| 0-5 | 444 (30%) | 414 (29%) | | 30 (45%) |  |
| 6-10 | 155 (10%) | 150 (11%) | | 5 (7.6%) |  |
| 11-20 | 315 (21%) | 300 (21%) | | 15 (23%) |  |
| 21-30 | 203 (14%) | 195 (14%) | | 8 (12%) |  |
| 31-40 | 130 (8.7%) | 127 (8.9%) | | 3 (4.5%) |  |
| 41-50 | 81 (5.5%) | 79 (5.6%) | | 2 (3.0%) |  |
| >50 | 158 (11%) | 155 (11%) | | 3 (4.5%) |  |
| Facility |  |  | |  | 0.042 |
| Kilifi County Hospital | 448 (30%) | 427 (30%) | | 21 (32%) |  |
| Matsangoni | 185 (12%) | 181 (13%) | | 4 (6.1%) |  |
| Mavueni | 312 (21%) | 296 (21%) | | 16 (24%) |  |
| Mtondia | 199 (13%) | 196 (14%) | | 3 (4.5%) |  |
| Pingilikani | 342 (23%) | 320 (23%) | | 22 (33%) |  |
| **Pediatric in-patient pneumonia surveillance** | | | | | |
| **Characteristic** | **Overall N = 391** | **Negative N = 327** | | **Positive  N = 64** | **p-value** |
| Sex |  |  | |  | 0.12 |
| Female | 179 (46%) | 144 (44%) | | 35 (55%) |  |
| Male | 212 (54%) | 183 (56%) | | 29 (45%) |  |
| Median Age (Months) | 11 (8, 17) | 11 (8, 17) | | 11 (7, 21) | >0.9 |
| Age Category (Months) |  |  | |  | 0.015 |
| 0–5 | 52 (13%) | 40 (12%) | | 12 (19%) |  |
| 6–11 | 147 (38%) | 124 (38%) | | 23 (36%) |  |
| 12–23 | 138 (35%) | 124 (38%) | | 14 (22%) |  |
| 24–59 | 38 (9.7%) | 26 (8.0%) | | 12 (19%) |  |
| >60 | 16 (4.1%) | 13 (4.0%) | | 3 (4.7%) |  |
| **In-patient diarrhea etiology surveillance** | | | | | |
| **Characteristic** | **Overall  N = 1,088** | **negative N = 1,040** | | **positive N = 48** | **p-value** |
| Sex |  |  | |  | 0.3 |
| Female | 477 (44%) | 454 (44%) | | 23 (52%) |  |
| Male | 604 (56%) | 583 (56%) | | 21 (48%) |  |
| Unknown | 7 | 3 | | 4 |  |
| Median Age (Months) | 6 (1, 12) | 5 (0, 12) | | 9 (4, 19) | 0.007 |
| Unknown | 7 | 3 | | 4 |  |
| Age Category (Months) |  |  | |  | 0.5 |
| 0–5 | 538 (50%) | 521 (50%) | | 17 (39%) |  |
| 6–11 | 244 (23%) | 233 (22%) | | 11 (25%) |  |
| 12–23 | 173 (16%) | 164 (16%) | | 9 (20%) |  |
| 24–59 | 126 (12%) | 119 (11%) | | 7 (16%) |  |
| Unknown | 7 | 3 | | 4 |  |

**Supplementary Figures**

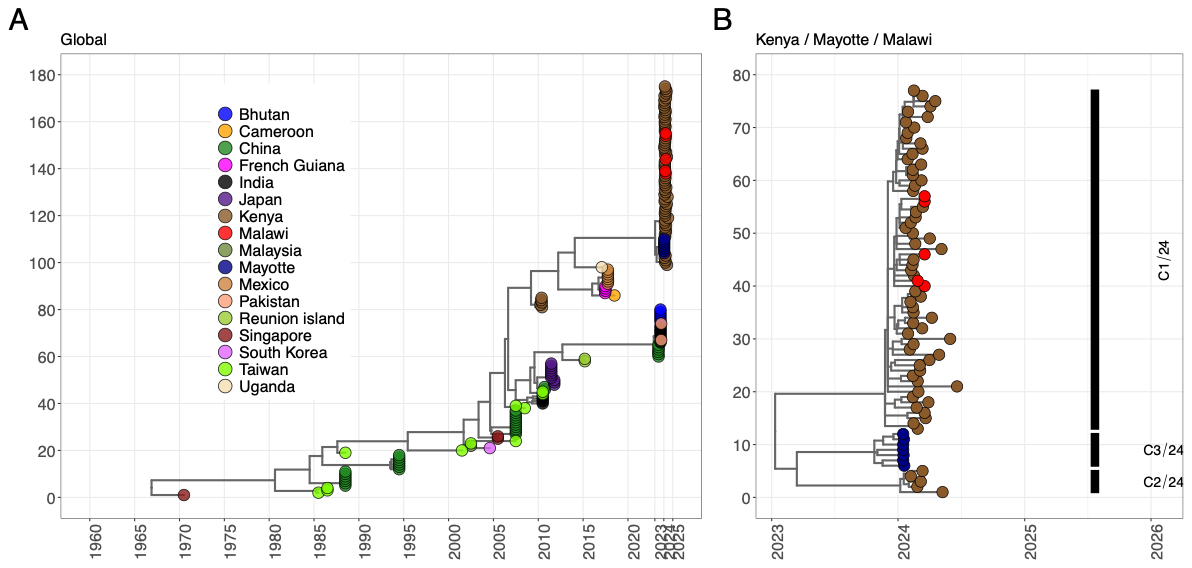

**Supplementary Figure 1.** Time-scaled phylogenetic tree showing the evolutionary placement of Kenyan CV-A24v sequences within the global CV-A24v diversity. (**A**) Global CV-A24v phylogeny. (**B**) Kenyan, Malawi and Mayotte sequences shown as an extension of the global tree. Tip points are colored according to the country of collection. Major clades within the East African region are labelled C1/24, C2/24 and C3/24 to highlight their phylogenetic grouping/clustering.

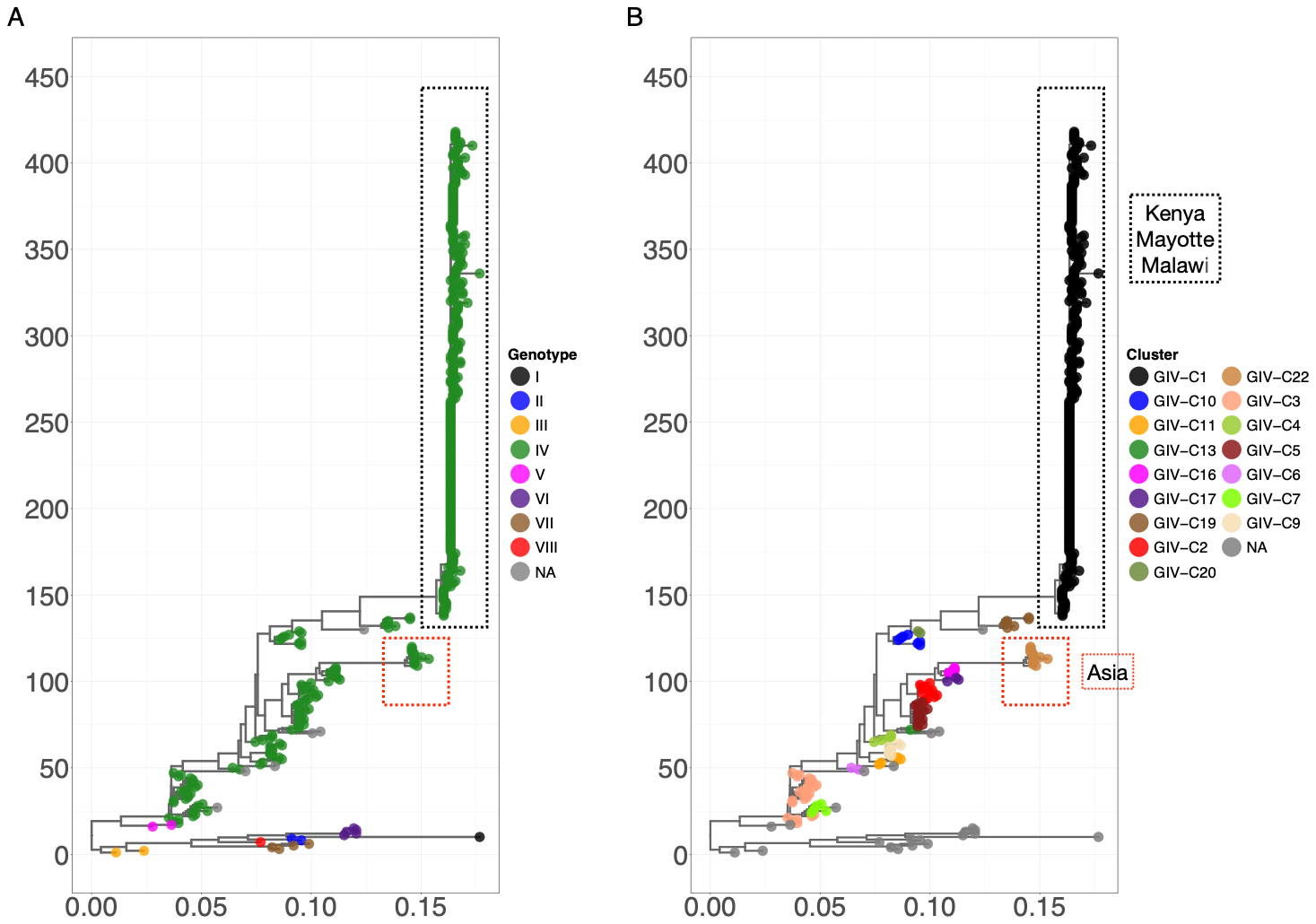

**Supplementary Figure 2**. Genetic diversity of globally circulating CV-A24v.

**(A)** ML phylogenetic tree of global CV-A24v VP1 sequences sampled between 1970 and 2024, showing the clustering of major CV-A24v genotypes. **(B)** ML phylogenetic tree of genotype IV CV-A24v VP1 sequences, showing clustering into distinct sub-groups based using a 0.05 nucleotide divergence threshold.

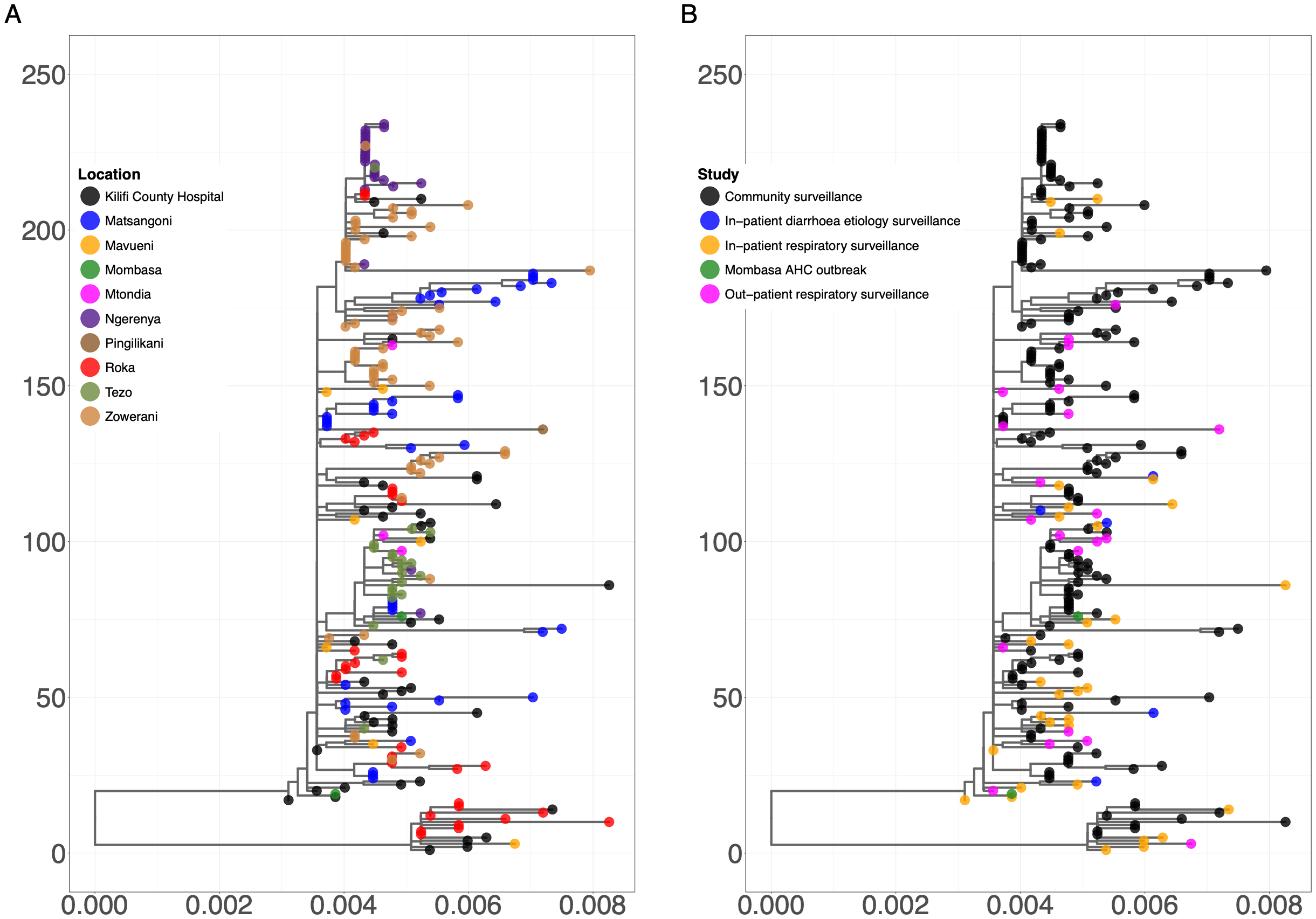

**Supplementary Figure 3.** ML phylogenetic trees of CV-A24v strains sampled during the coastal Kenya the 2024 AHC outbreak (n=245). (**A**) Tip labels are colored by sampling administrative location within Kilifi county. (**B**) Tip labels are colored by surveillance platforms.

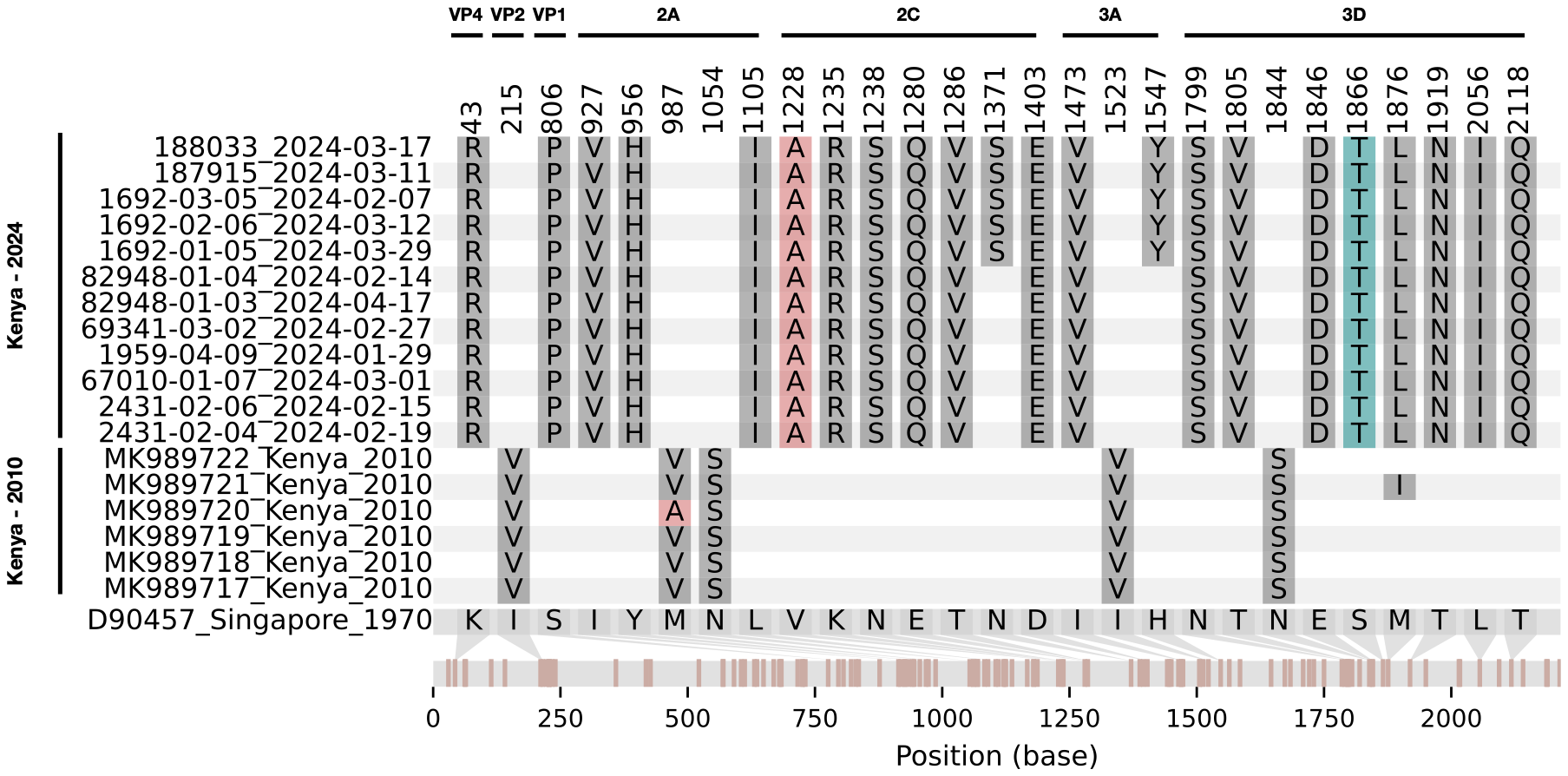

**Supplementary Figure 4**. Amino acid variability in CV-A24v genomes from the Kenyan 2024 and 2010 outbreaks.

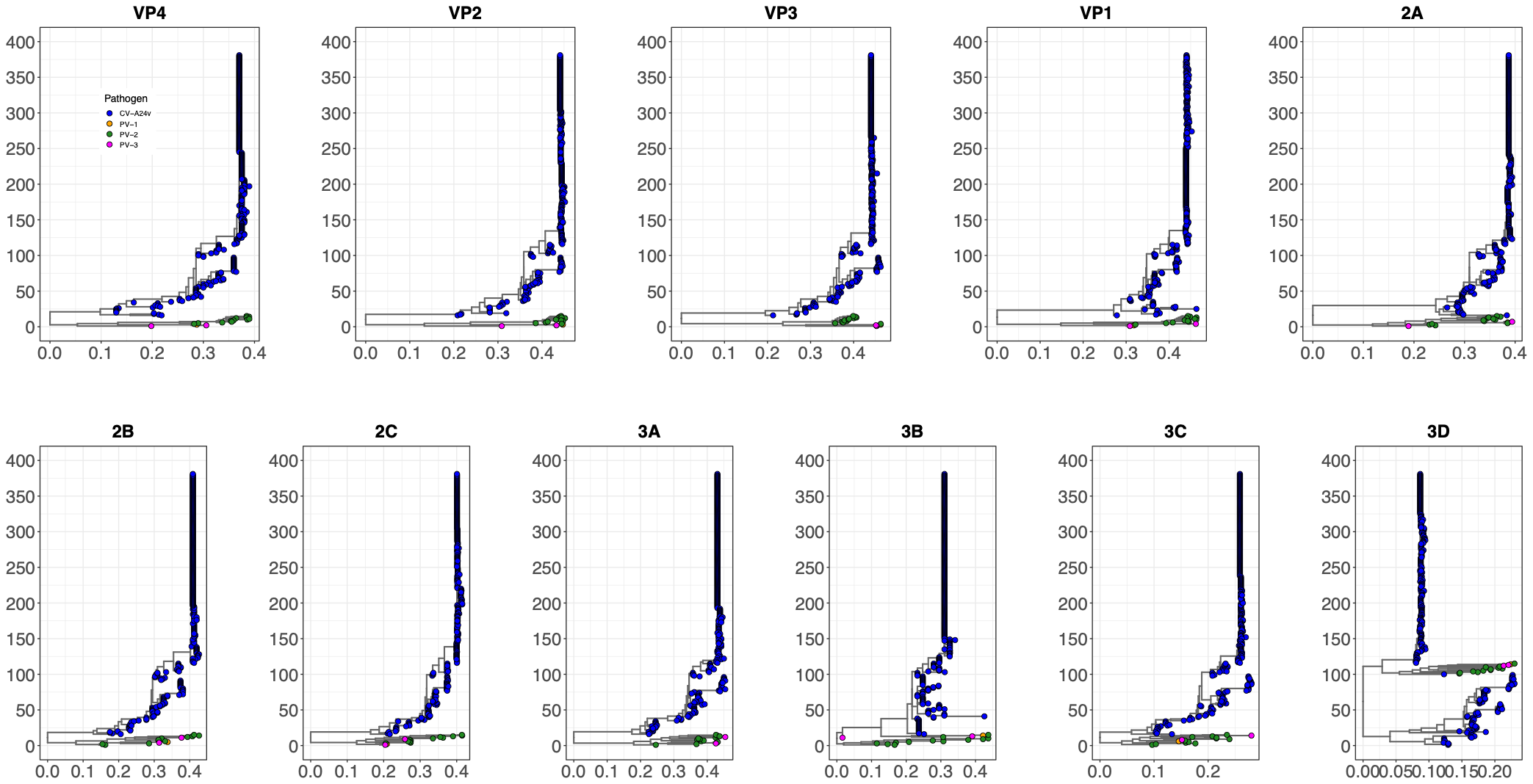

**Supplementary Figure 5.** ML phylogenetic trees for individual coding regions of CV-A24v and suspected recombinant viruses. Tip labels colors represent pathogen.

**CV-A24 detection in the various surveillance platforms**

- In the ResViRe study, CV-A24v was detected in 375 samples by RT-PCR and VP4/VP2 sequencing; 188 samples by RT-PCR, VP4/VP2 sequencing, and whole-genome sequencing; 31 samples by RT-PCR alone; and 3 samples by RT-PCR with whole-genome sequencing.
- In the outpatient ARI study, CV-A24v was detected in 257 samples RT-PCR alone, and 22 samples by RT-PCR with whole-genome sequencing.
- In the inpatient pediatric pneumonia study, 12 samples were detected by RT-PCR alone, and 32 samples by RT-PCR with whole-genome sequencing.
- In the pediatric inpatient diarrhoea surveillance, 57 samples were detected by RT-PCR, and 5 samples by RT-PCR with whole-genome sequencing.
